# Positive Mental Training in patients with Multiple Sclerosis experiencing psychological distress: A Preliminary Randomised Controlled Trial

**DOI:** 10.1101/2021.07.15.21260604

**Authors:** Katy Murray, Giulia Melchiorre, Alastair Dobbin, Killian A Welch

## Abstract

**Introduction:** Psychological distress is a major issue in multiple sclerosis (MS), having a significant impact on quality of life. Antidepressants are generally unhelpful for subsyndromal symptomatology, and psychological treatment approaches often not accessible or too cognitively demanding for some patients. There is an urgent need for low-cost interventions to improve wellbeing in MS.

**Methods:** This was a pilot randomised controlled trial (RCT) of Positive Mental Training (PosMT), a low intensity intervention providing training in positivity, optimism and resilience previously shown to improve anxious and depressive symptomotology. 28 patients with MS were randomised to the intervention and 30 to the control group.

**Results:** Follow-up data was obtained from 39 patents. The majority of participants receiving PosMT reported that they had used the intervention, with few reporting side effects. The intervention group reported a significant improvement in self-rated health as measured by the EuroQual visual analogue scale, F(4,34) = 3.204, p = 0.025, R2 = 0.274.

**Discussion:** This preliminary RCT found that PosMT in its current form could be used by patients with MS with little difficulty. Despite the small size of the study, allocation to the intervention was found to be associated with a significant improvement in self-rated health. Given the low cost of PosMT and its easy availability (it can simply be downloaded from a website), this pilot RCT suggests it could be a useful tool for MS patients. We believe this intervention warrants further study, ideally in a large multi-centre RCT.

## Introduction

Psychological distress is common in multiple sclerosis (MS),(Boeschoten RE *et al*) impacts adversely on quality of life,(McGuigan C and Hutchinson M) and goes largely untreated Cetin K *et al*). While antidepressants can treat depression in neurological conditions,(Price A *et al*) they are not acceptable to some patients and can be associated with side effects. Cognitive Behavioural Therapy (CBT) has reasonable evidence for treating depression in MS.(Hind D *et al*) It can be cognitively demanding however, necessitating relatively intact memory and executive function. Given that cognitive impairment is common in MS,(Amato MP) it may not be appropriate for many patients. CBT, as traditionally delivered by a highly trained paractioner on a 1:1 basis, is also an expensive intervention, this hampering delivery on a large scale basis. Indeed, there are doubts about its cost effectiveness when delivered to patients with relatively low levels of psychological distress (Mosweu *et al*). Positive Mental Training (PosMT) is a guided self-help treatment providing training in positivity, optimism and resilience. It improves anxiety and depression in primary care,(Dobbin A) but efficacy in MS has not been examined. We conducted a preliminary randomized controlled study to explore the tolerability and potential benefits of PosMT in MS patients experiencing some degree of psychological distress.

## Methods

PosMT consists of a training video and 12 audio tracks each lasting 18 minutes. It incorporates therapeutic techniques from relaxation, mindfulness and positive psychology. This was a single-centre trial, with patients randomised to twelve weeks of either intervention or standard treatment. After the twelve weeks, on receipt of follow-up ratings, the control group were provided with the intervention. The study protocol was approved by Lothian Ethics Committee and informed consent obtained from all participants. The study was registered at www.clinicaltrials.gov (NCT02524093).

Participants were recruited from MS clinics at the Anne Rowling Regenerative Neurology Clinic, Edinburgh. As PosMT would most likely benefit people experiencing some psychological distress, patients had to score more than 4 on either the Hospital Anxiety (HADS-A) or Depression Scale (HADS-D). In standard practice a cut of 8 is generally used to identify ‘ caseness’. (Honarmand K and Feinstein A) We felt a lower cut off was warranted, there being much ‘ subclinical’ psychiatric symptomatology in neurological disorders which can impact significantly on disability.(Carson AJ *et al*)

Participants were randomised in a ratio of 1:1 between PosMT or usual care. Usual care in Edinburgh consists of standard primary care input, annual Neurology follow up and access to a specialist MS nurse. Randomisation was through a computer generated sequence held on a webserver hosted within the University of Edinburgh, the allocation sequence concealed from study investigators.

Patients randomised to active treatment were provided with the intervention delivered by their preferred media (DVD/CDs, an mp3 download or an app). Patients randomised to standard care were sent a letter informing them they had been randomised to the control group and would be offered the intervention in 12 weeks, on returning repeat questionnaires. After 12 weeks all participants were posted the follow up questionnaires and asked to return them, the latter group being provided with the programme on receipt of completed questionnaires. Participants who did not return questionnaires received a telephone prompt to return the forms from either KM or KW. If contact was not made, or questionnaires not received within two weeks, they were posted a reminder letter and further set of forms. Given the nature of the study, patients obviously were not blinded to group allocation.

### Outcome measures

A priority was to ascertain recruitment, utilisation and drop-out rates and acceptability of the intervention to patients with MS. Adverse events in both groups were recorded throughout the trial. In addition, at the twelve-week assessment point, we asked those receiving the active treatment to complete a questionnaire exploring their experience of PosMT and any problems encountered with it.

This was a preliminary study, the primary aim of which was to establish the acceptability of PosMT to patients with MS and their ability to use the intervention without modifications. Nonetheless, we hoped that a reasonable trial size of 60 participants (30 randomised to intervention and 30 to control) may provide some indication of efficacy of the intervention in this population. The outcome measures selected were changes in ratings on the Hospital Anxiety and Depression Scale (HADS; to assess symptoms of anxiety and depression), Short Warwick-Edinburgh Mental Well-being Scale (SWEMWBS, a measure of general wellbeing) and the EuroQol health related quality of life -5 dimensions -5 levels (EQ-5D, a measure of health-related quality of life). These measures were chosen because of their widely accepted utility in measuring psychological distress (HADS), established ability to monitor wellbeing (SWEMWB) and utility in measuring the benefits of different interventions on a common simple metric and ubiquitous use in health economics research (EQ-5D). Due to an administrative error, the HADS was not sent out at the second time point so we do not have post-intervention data for this measure.

### Statistical analysis

Statistical analyses were performed using the SPSS for Windows statistical software package (version 22.0; SPSS, Inc, Chicago, IL). Baseline demographics between groups were compared using Student’ s t-tests or Mann-Whitney U tests for interval data (if normally or non-normally distributed respectively), or the Chi-squared test for category data. The effect of the intervention was interrogated by running a multiple linear regression model to determine whether positive mental training affected the change in the EQ5index score, the EQVAS or the SWBMES. This approach was necessary because of baseline differences between the intervention and control group on certain wellbeing and quality of life measures and because some of the outcome data was not normally distributed (Shapiro-Wilk p > 0.05). Only patients with baseline and follow up data were included in these analyses; patients randomised to the intervention with follow up data were included regardless of whether they reported having made use of PosMT. Further details of the statistical model are available in supplementary data (S1).

## Results

58 patients were recruited. 28 were randomised to intervention and 30 to treatment as usual. Participant flow is detailed in Figure 1 in the supplementary data. There were statistically significant baseline differences between intervention and control groups some measures (see Table 1), controlled for in the multiple linear regression model.

**Table 1:** Baseline assessments

|  | All randomised participants |  |  | Participants with follow up data |  |  |
| --- | --- | --- | --- | --- | --- | --- |
|  | Intervention | Control | Statistical comparison | Intervention | Control | Statistical comparison |
| **Total number** | 28 | 30 | Chi Square | 20 | 19 | Chi Square |
| **Male** | 6 | 7 | X(1) = 0.030,<br>p = 0.862 | 5 | 4 | X(1) = 0.086,<br>p = 0.770 |
| **Female** | 22 | 23 |  | 15 | 15 |  |
| **Age (SD)** | 43.43 (10.45) | 45.07 (11.43) | T-test<br>t(56) = -0.568,<br>p = 0.572 | 44.80 (10.11) | 41.58 (12.07) | T-test<br>t(37) = 0.905,<br>p = 0.371 |
| **Range** | 26 to 66 | 20 to 66 |  | 31 to 66 | 20 to 66 |  |
| **Illness duration (SD)** | 6.96 (5.63) | 8.30 (7.23) | Mann Whitney U<br>U = 398.5, p = 0.737 | 7.08 (5.71) | 7.32 (6.88) | Mann Whitney U<br>U = 180.0,<br>p = 0.778 |
| **Range** | 1 to 22 | 1 to 23 |  | 1 to 22 | 1 to 23 | - |
| **HADS Total (SD)** | 14.79 (5.16) | 17.00 (6.33) | T-test<br>t(56) = -1.454 ,<br>p=0.152 | 14.60 (5.45) | 16.21 (5.77) | Mann Whitney U:<br>U = 164.5,<br>p=0.472 |
| **HADS-A (SD)** | 8.86 (3.83) | 10.50 (4.10) | t(56) = -1.575 ,<br>p=0.121 | 8.45 (4.07) | 10.53 (3.91) | Mann Whitney U:<br>U = 123.5,<br>p = 0.060 |
| **HADS-D (SD)** | 5.93 (3.95) | 6.50 (4.02) | t(56) = -0.545 ,<br>p=0.588 | 6.15 (4.07) | 5.68 (3.37) | T-test:<br>t(37) = 0.388,<br>p=0.700 |
| **SWEMWBS (SD)** | 21.21 (3.02) | 19.57 (3.05) | T-test<br>t(55) = 2.039,<br>p = 0.046* | 21.42 (3.20) | 19.36 (2.42) | T-test:<br>t(37) = 2.265,<br>p = 0.029* |
| **EuroQol-5D-5L Index (SD)** | 0.61 (0.21) | 0.66 (0.20) | Mann Whitney U<br>U= 349.5,<br>p = 0.272 | 0.59 (0.20) | 0.71 (0.18) | Mann Whitney:<br>U = 112.0,<br>p = 0.028* |
| **EuroQol VAS (SD)** | 67.00 (12.73) | 69.37 (15.71) | T-test<br>T(56) = - 0.627,<br>p = 0.533 | 65.40 (10.59) | 72.16 (15.25) | t-test:<br>t(37)=-1.615,<br>p = 0.115 |

\*Indicates statistically significant difference between groups ( P < 0.05)

**Figure 1.**
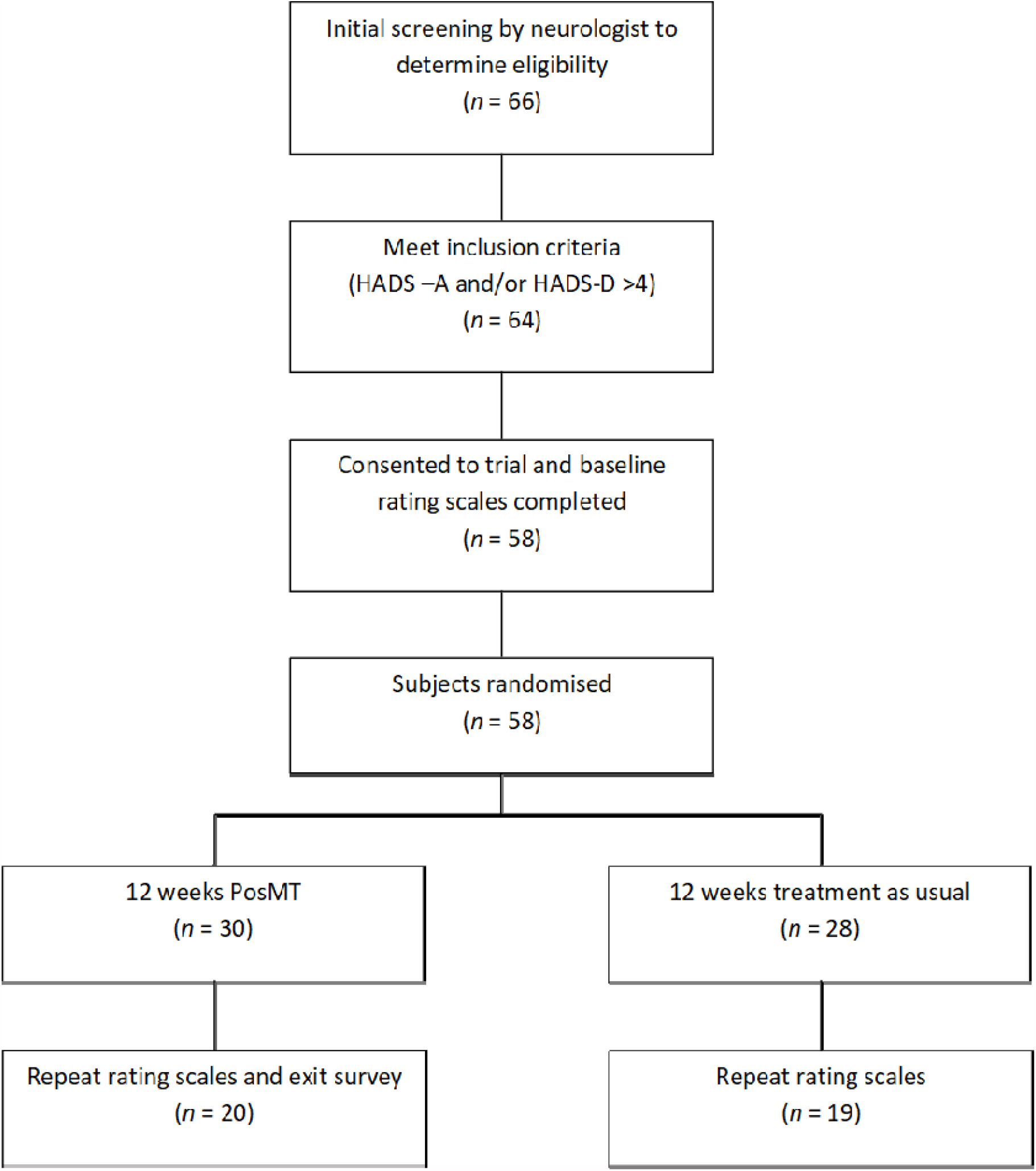
Flow of patients through trial

Full follow up data was obtained for 37 (63.79%) and full or partial follow up data for 39 (67.24%) participants. Drop out analysis revealed no differences in response rates based on age, gender, illness duration, baseline HADS rating or group assignment (p > 0.05, see supplementary data).

A multiple linear regression was run to predict changes in scores for the EuroQol-5D-5L index, EuroQol VAS, and SWEMWBS over 12 weeks from group allocation (treatment vs control), gender, illness duration, and baseline total HADS score (Table 2). The variables statistically significantly predicted change in EuroQol VAS, F(4,34) = 3.204, p = 0.025, R2 = 0.274. Only group allocation added significantly to the prediction, p = 0.007. Change in EQ VAS rating between baseline and 12 weeks is shown graphically in Figure 2. No significant effect was seen on ratings on the other two outcome measures. The full model for the EuroQal VAS analysis is available in the supplementary data (S2).

**Table 2.**
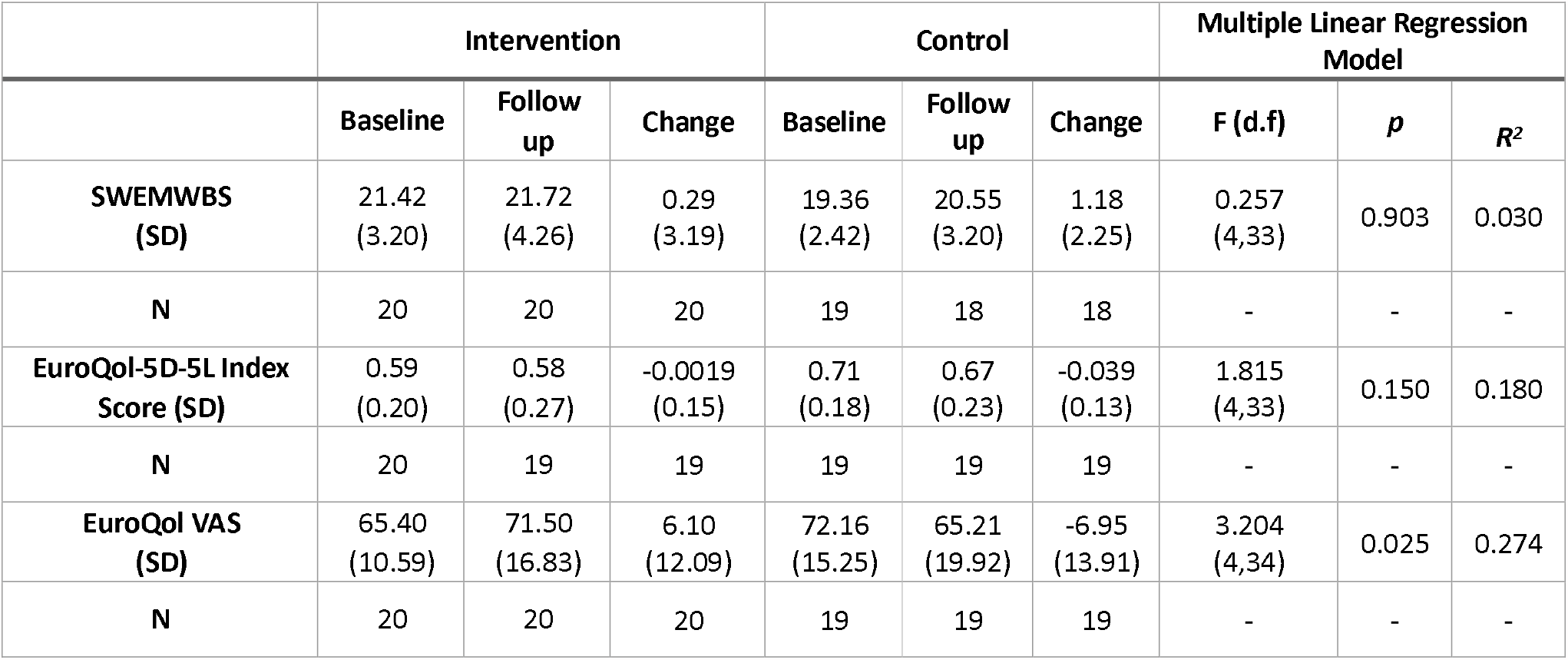
Comparison of change in score on rating scales after 12 weeks in intervention and control groups.

|  | Intervention |  |  | Control |  |  | Multiple Linear Regression Model |  |  |
| --- | --- | --- | --- | --- | --- | --- | --- | --- | --- |
|  | Baseline | Follow up | Change | Baseline | Follow up | Change | F (d.f) | *p* | *R*<sup>2</sup> |
| **SWEMWBS (SD)** | 21.42<br>(3.20) | 21.72<br>(4.26) | 0.29<br>(3.19) | 19.36<br>(2.42) | 20.55<br>(3.20) | 1.18<br>(2.25) | 0.257<br>(4,33) | 0.903 | 0.030 |
| **N** | 20 | 20 | 20 | 19 | 18 | 18 | - | - | - |
| **EuroQol-5D-5L Index Score (SD)** | 0.59<br>(0.20) | 0.58<br>(0.27) | -0.0019<br>(0.15) | 0.71<br>(0.18) | 0.67<br>(0.23) | -0.039<br>(0.13) | 1.815<br>(4,33) | 0.150 | 0.180 |
| **N** | 20 | 19 | 19 | 19 | 19 | 19 | - | - | - |
| **EuroQol VAS (SD)** | 65.40<br>(10.59) | 71.50<br>(16.83) | 6.10<br>(12.09) | 72.16<br>(15.25) | 65.21<br>(19.92) | -6.95<br>(13.91) | 3.204<br>(4,34) | 0.025 | 0.274 |
| **N** | 20 | 20 | 20 | 19 | 19 | 19 | - | - | - |

**Figure 2.**
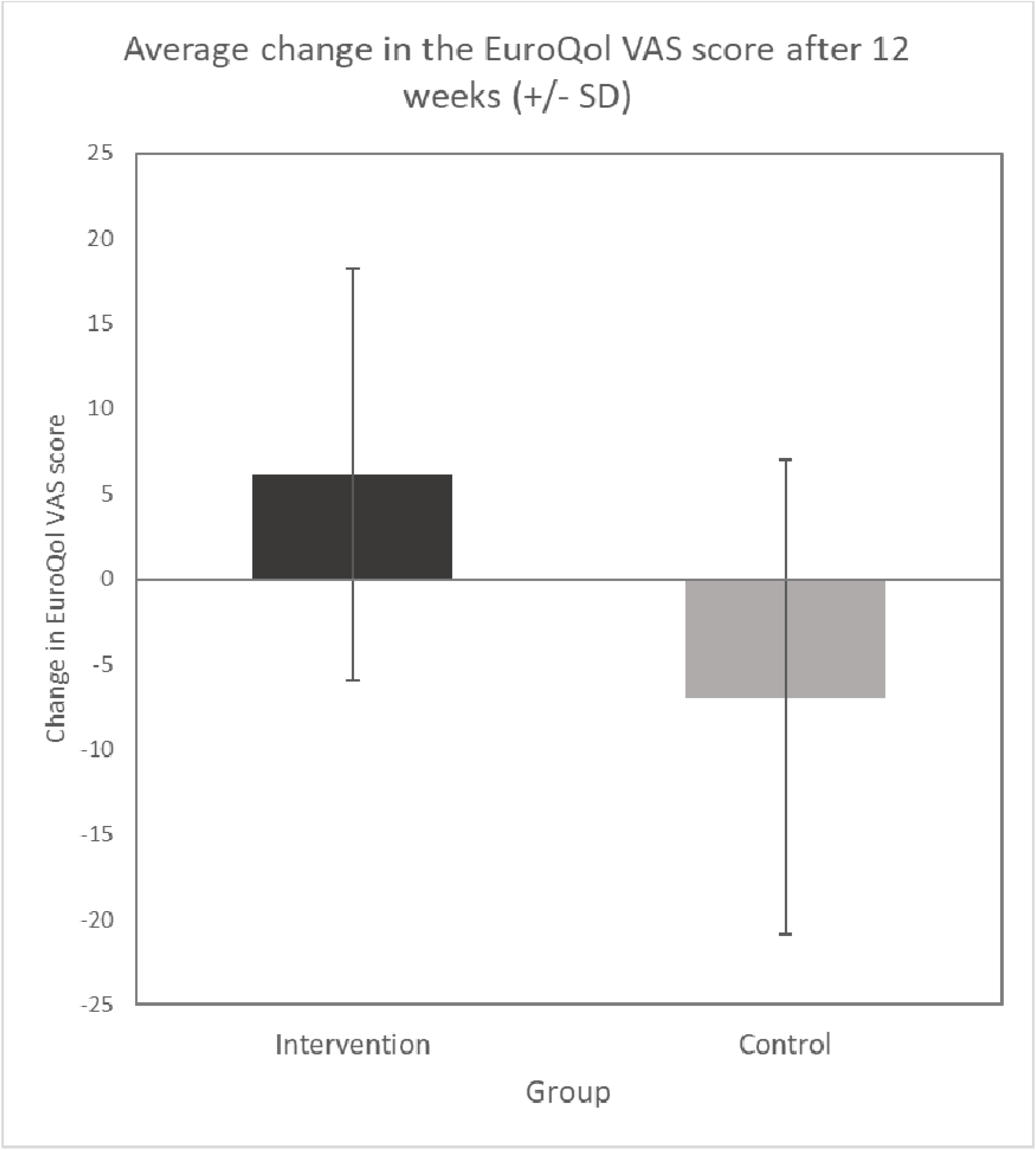
Graphical depiction of mean change in the EuroQol VAS score in intervention and control groups after 12 weeks.

Seventeen of the twenty PosMT patients providing repeat rating scales also returned forms exploring their experiences of the intervention. This is further discussed in supplementary data, but revealed that most listened to all tracks and the intervention was well tolerated. There were few suggestions for improvements.

## Discussion

This preliminary study suggests that PosMT is accessible by and acceptable to patients with MS. It was associated with significant improvements in self-rated health on one measure (the EuroQal VAS), with qualitative data indicating that several patients found the intervention very helpful. Overall it was well tolerated, with minimal side effects. While there are issues with the study which require further discussion, given the low cost and low intensity of the intervention these findings potentially have considerable clinical relevance.

A finding that requires consideration is that while PosMT was associated with significant improvement in self-rated health as measured by the EuroQol VAS, no change was seen in the EuroQol Index, or wellbeing as measured by the SWEMWEBS. While this could simply be a spurious finding, a possible explanation is that the EuroQol VAS does yield a truly ‘ global’ measure of self-rated health, whereas the EuroQol Index score is simply an amalgamation of ratings on each of the five dimensions (and consequently doesn’ t capture all aspects of self-perceived health). Inkeeping with this, it is recognised that EuroQol-VAS does not always strongly correlate with EuroQol Index scores.(Feng Y *et al*)

There are several limitations of the study. Firstly, we had intended to also examine whether PosMT was associated with a change in anxiety and depressive symptoms, as measured by the HADS. As discussed however, follow up HADS ratings were not obtained. While we did explore if the intervention was associated with change in the ‘ anxiety/depression’ dimension of the EuroQual (and no such effect was seen), we acknowledge that this is likely to be a crude and insensitive measure of anxious and depressive symptomatology. It certainly does not enable us to dismiss the possibility that the improvements in self-perceived health observed in association with the intervention are driven by improvements in these symptoms. What is notable however is that baseline HADS score did not influence findings. This was surprising, as one would expect that people with greater psychopathology would be more likely to benefit from PosMT. Indeed, this has been previously reported.(Koeser L *et al*) It may be that our study numbers were too small to observe a statistically significant finding. It does not seem likely that our patient group simply didn’ t exhibit enough psychopathology to enable such an effect to be mediated. In keeping with the known high rates of pychopathology in this patient group, the mean total HADS score was 15.7; a score of 8 in either the anxiety or depression components is generally taken as ‘ caseness’. (Honarmand K *et al*)

It is also the case that follow up data was obtained from fewer participants than we hoped, (67% randomised to intervention and 63% of controls). Though not ideal, this does actually compare favourably with other trials of low-intensity interventions.(Andersson G *et al* and Richards D *et al*) Additionally, the intervention group rated wellbeing as higher and quality of life lower than controls at baseline. It seems highly unlikely this drove the findings however. Aside from the fact that the baseline differences between the two groups on EQ VAS rating were not significantly different (this being the measure in which a significant intervention effect was seen), the approach to statistical analyses controlled for any baseline differences present. Presumably the fact that these baseline differences were present simply reflects the small size of the study.

Lastly, we did employ an active control. While we did consider this, given the low intensity of the intervention it was difficult to identify a comparator which would neither overlap considerably with the intervention itself (e.g. a relaxation package) or potentially antagonise patients with its low ‘ face’ efficacy (e.g. music CDs). On balance we felt that, given this was a preliminary study, a ‘ treatment as usual’ control was reasonable.

In summary therefore, while acknowledging our findings are preliminary, this study suggests that PosMT has potential in treating psychological morbidity in MS. It is cheap, acceptable to patients, and generally well tolerated. Further examination of its efficacy in MS seems warranted.

## Data Availability

Supplementary data included

## Supplementary data

### Drop out analysis

- Assigned each participant to one of to groups: drop-out yes vs drop-out no
- Included patients in this analysis that were not assigned to either of the two groups
- Checked for significant differences in baseline variables, including age, gender, illness duration, baseline HADS rating or group assignment

**Table 3.** drop out analysis details

|  | Dropped out | Did not drop out | Statistical comparison |
| --- | --- | --- | --- |
| **Total number** | 25 | 39 |  |
| **Male** | 5 | 9 | Chi Square<br>$\chi^2(1) = 0.084, p = 0.771$ |
| **Female** | 20 | 30 |  |
| **Age (SD)** | 45.72 (10.56) | 43.23 (11.08) | T-test $t(62) = 0.893, p = 0.375$ |
| **Range** | 25 to 62 | 20 to 66 |  |
| **Illness duration (SD)** | 8.92 (7.45) | 7.19 (6.22) | Mann Whitney U<br>$U = 398.5, p = 0.737$ |
| **Range** | 1 to 24 | 1 to 23 |  |
| **HADS Total (SD)** | 16.83 (6.25) | 15.38 (5.59) | Mann Whitney U<br>$U = 335.5, p = 0.187$ |

### Full model below for EuroQol VAS

| Coefficients<sup>a</sup> |  |  |  |  |  |  |  |  |
| --- | --- | --- | --- | --- | --- | --- | --- | --- |
| Model |  | Unstandardized Coefficients |  | Standardized Coefficients | t | Sig. | Collinearity Statistics |  |
|  |  | B | Std. Error | Beta |  |  | Tolerance | VIF |
| 1 | (Constant) | 25.567 | 11.325 |  | 2.258 | .031 |  |  |
|  | Gender | 1.314 | 5.039 | .039 | .261 | .796 | .962 | 1.040 |
|  | Group | -12.014 | 4.212 | -.421 | -2.852 | .007 | .978 | 1.022 |
|  | HADS Total Baseline | -.677 | .397 | -.262 | -1.702 | .098 | .900 | 1.111 |
|  | Illness duration (Years) | .018 | .347 | .008 | .051 | .959 | .953 | 1.050 |

a. Dependent Variable: Change\_EQVAS

## Findings from questionnaire (qualitative data)

Of the 17 patients who returned completed questionnaires exploring their experience of the intervention all listened to at least 2 of the tracks. Indeed, 11 listened to all 12 and the mean number of tracks listened to was 9.8. Sixteen of these patients estimated the total numbers of tracks listened to, with a range of 6 to 84 and mean of 53.7. Of the twelve who reported at least one track they found most helpful (with many patients citing more than one track), these were track one (9), track two (7), track three (3), track four (2), track five (2), track eight (1) and ‘ all’ (1, not included in preceding numbers). At least one least helpful track was reported by ten patients. This was track two (1), three or four (3), eight (2), nine (3), ten (1), eleven (1) and twelve (1). Ten made response when asked how the intervention could be improved. Five responded that they could think of no improvements, often expanding that it had been very helpful; two found the music annoying; two found the voice annoying; one thought it too sports focussed; one stated fist-clenching brought on MS symptoms; and one felt the intervention should be paired with counselling or therapy. Participants were also asked if the intervention had been associated with side effects. As well as the report from one participant that fist-clenching brought on MS symptoms, one reported that twenty minutes a day was a lot of time, and two that they fell asleep when listening.

